# Incorporating Imaging Markers of Brain Health in Modeling of Functional Outcome After Acute Ischemic Stroke: A Quantitative Comparison Study

**DOI:** 10.1101/2025.05.02.25326881

**Authors:** Erik Lindgren, Luca Angeleri, Martin Bretzner, Anna K. Bonkhoff, Christina Jern, Arne G Lindgren, Jane Maguire, Robert W. Regenhardt, Natalia S. Rost, Markus D. Schirmer, the MRI-GENIE and GISCOME Investigators

## Abstract

**Background and Objectives:** Brain health facilitates resilience to withstand detrimental effects of brain disease, but is challenging to assess in the time-sensitive setting of acute ischemic stroke (AIS). In this international observational multicenter study, we compare quantitative MRI markers of structural brain health in their ability to predict functional outcome after AIS.

**Methods:** We included AIS patients from the international MRI-GENIE study (multicenter; 2003-2011) with acute T2-FLAIR imaging. Automated pipelines estimated white matter hyperintensity volume (WMHv), brain volume, and intracranial volume (ICV). We normalized WMHv by individual brain volume, creating WMH load for each patient.

Assessed brain health markers included: brain parenchymal fraction (brain volume relative to ICV); radiomics derived brain age (based on an ElasticNet linear regression model); brain reserve (normal appearing brain volume [brain volume minus WMHv] relative to ICV), and effective Reserve (eR, latent variable based on age, WMH load [WMHv and brain volume). We added the markers to a clinical reference model (including age, sex, diabetes type 2, hypertension, NIHSS, atrial fibrillation and prior stroke), comparing model performances between separate multivariable regression models in their prediction of unfavorable outcome (90-day modified Rankin Scale score 3-6), using Bayesian Information Criterion (BIC).

**Results:** We analyzed 2,303 patients (median age 66 years, 46% female, 27% unfavorable outcome) after excluding 357/2,660 patients (13.4%) due to missing follow-up data. Comparison using BIC provided strong statistical evidence (ΔBIC > 6) for all four models using a brain health marker to outperform the clinical reference model (BIC=2354.9). The eR model showed the lowest BIC value (BIC=2318.4), providing very strong (ΔBIC > 10) statistical evidence to outperform all other models. The model utilizing radiomics derived brain age showed the second lowest BIC value (BIC=2329.3), providing very strong (ΔBIC > 10) statistical evidence to outperform all other models except the eR model.

**Discussion:** Incorporating quantitative MRI markers of brain health in predictive models enhance personalized outcome prognostication after AIS. Our results suggest eR as a viable marker for future studies investigating structural brain health and ischemic stroke.

## Introduction

Brain health facilitates resilience to withstand detrimental effects of brain disease, and is a prioritized research area by international organizations.^1^ As a holistic and evolving concept, brain health encompasses both absence of disease and the brain’s optimal functioning during a lifespan, even if a disease is present.^1,2^ Preserving and understanding threats to brain health is essential for preventing and enhancing resilience against age-related neuropathology. ^3^

While stroke is the leading global contributor to long-term disability, the importance of brain health for functional outcome after acute ischemic stroke (AIS) remains relatively underexplored.^4^ However, recently developed automated evaluation of magnetic resonance imaging (MRI) markers at time of admission presents opportunities for prompt quantification of structural brain health markers, with the potential to inform personalized clinical decision making even in acute settings. The purpose with this study was to investigate whether incorporation of MRI-markers of structural brain health improve modeling of functional outcome after acute ischemic stroke.

We aimed to compare four quantitative MRI markers of brain health that can be assessed in the acute stroke setting using automated deep learning enabled pipelines and routinely obtained MRI sequences. In a large multicenter clinical cohort, we compared these markers to each other and to a reference model with only clinical variables in terms of the ability to predict functional outcome after AIS.

## Methods

This report adheres to international guidelines for reporting observational studies.^5^

### Standard protocol approval, registration and patient consent

Participating centers received permission from local authorities and ethics committees to collect observational data and obtained written informed consent from participants or their surrogates at time of enrollment, when required under applicable laws.

### Participants

We analyzed data from the MRI-GENetics Interface Exploration (MRI-GENIE) study, an international AIS cohort (multicenter, 2003-2011). Details of the MRI-GENIE cohort have been described previously.^6^ Patients with acute MRI including T2-Fluid-attenuated inversion recovery (FLAIR) sequences were eligible. Participants aged 90 years and older were aggregated into a single age category in accordance with the Health Insurance Portability and Accountability Act (HIPAA).

### Phenotypical data

Clinical characteristics (stroke severity according to National Institutes of Health Stroke Scale [NIHSS]), demographics and medical history were collected at acute hospital admission. Functional outcome was assessed at approximately 90 days post-stroke, using the modified Rankin Scale (mRS), a 7-grade ordinal scale ranging from 0 representing no symptoms, to 6 representing death.^7^ We defined unfavorable outcome as mRS score 3-6.^8^

### Neuroimaging

Neuroimaging was obtained mostly within 48 hours of acute hospital admission (median 2 days, IQR 1-4 days) as part of routine care using AIS protocols. All images were either captured on 1T, 1.5T or 3T scanners (General Electric Medical Systems, Philips Medical Systems, Siemens, Toshiba, Marconi Medical Systems, Picker International Inc.). We automatically assessed white matter hyperintensity (WMH) volume, intracranial volume (ICV) and brain volume (combined gray matter and white matter parenchymal volumes) based on FLAIR, utilizing pipelines developed for MRI acquired in the acute clinical setting of AIS patients.^9,10^ Results were manually reviewed, and failed segmentations, e.g. due to image artifacts, were excluded. All volumes (WMH volume, brain volume) were calculated multiplying number of voxels by voxel size. We normalized WMH volume by individual brain volume, creating WMH load for each patient.

### MRI brain health markers

We assessed four neuroimaging markers of brain health described in the literature: 1) brain parenchymal fraction (BPF)^11^ – brain volume relative to ICV; 2) radiomics derived brain age (BA)^12^ – based on an ElasticNet linear regression model; 3) brain reserve (BR)^13^ – normal appearing brain volume (brain volume minus WMH volume) relative to ICV; and 4) effective Reserve (eR)^14^ – a latent variable estimated based on the observed variables age, WMH load and brain volume. To prevent overfitting, pathway coefficients used to estimate eR in the study cohort were derived from a separate cohort.^14^

### Statistical analysis

We report continuous data as medians with interquartile ranges (IQR) and dichotomous data as proportions of investigated patients. We assessed normal distributions using Shapiro-Wilk test and applied logit transformation on WMH load for normality. We excluded patients with incomplete data (see Figure 1). Number of included patients from each participating center appear in eTable 2.

**Figure 1.**
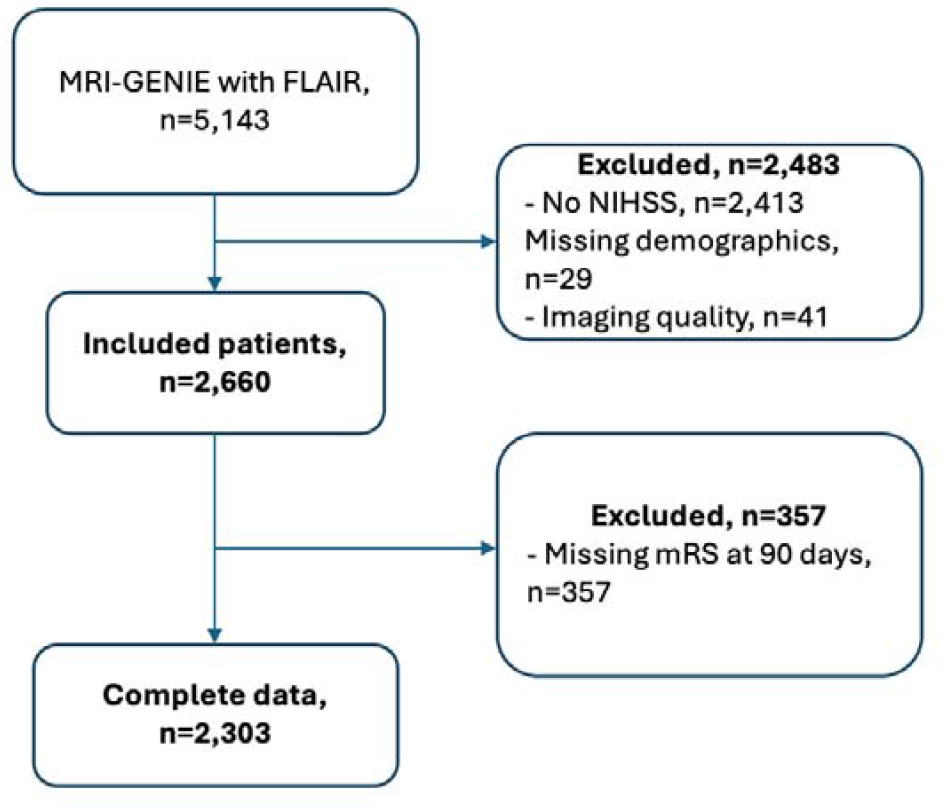
Flow Chart of Included Patients. Abbreviations: MRI-GENIE - MRI-GENetics Interface Exploration study; NIHSS – National Institutes of Health Stroke Scale; mRS – modified Rankin Scale

We defined a clinical reference logistic regression model based on biological plausibility and consistent reporting as independent predictors for unfavorable outcome (mRS score 3-6 at 90 days). The model included clinical variables such as age, sex, diabetes mellitus type 2, hypertension, atrial fibrillation, history of prior stroke and stroke severity (NIHSS). To this reference model, MRI brain health markers were added creating four brain health models while controlling for multicollinearity using variance inflation factor (VIF). To mitigate inclusion of highly correlated variables into the same model, adjustment for age was only conducted if not used to determine the respective brain health marker (see Table 2).

We compared the predictive performances between these four brain health models and the reference model in terms of goodness-of-fit using Bayesian Information Criterion (BIC).^15^ A lower BIC value represents relatively superior model performance, and the methodology involves penalizing to more complex models to minimize risk for overfitting. In model selection analysis, evidence levels for statistical differences are considered weak, positive, strong or very strong for differences in BIC (ΔBIC) values larger than 0, 2, 6 and 10 respectively.^15^ We further calculated the area under the precision-recall curve (PR-AUC) with 95% CI using 10-fold cross-validation with 100 iterations. In each iteration, test cohorts were formed by randomly selecting 100 positive and negative outcomes defined as mRS 3-6 and 0-2 respectively. The remaining dataset (full dataset - test cohort), were used to train the models. The PR-AUC resembles the traditional receiver operating characteristic AUC, while being more accurate in unbalanced datasets.^16^ As the outcome measured in this cohort was unbalanced, with approximately one fourth of the patients categorized as having an unfavorable outcome, PR-AUC was used instead of ROC-AUC. Mean PR-AUC values and corresponding 95% CI were calculated in the test cohorts from stored predicted probabilities of PR-AUC integrals. P-values were calculated using Mann Whitney U tests with significance level set to p<0.05.

In a subanalysis, we conducted the same analyses in more complex models, additionally including comprehensive adjustments also for WMH load and brain volume if not used to determine the respective brain health marker (see eTable 1).

**Table 1.**
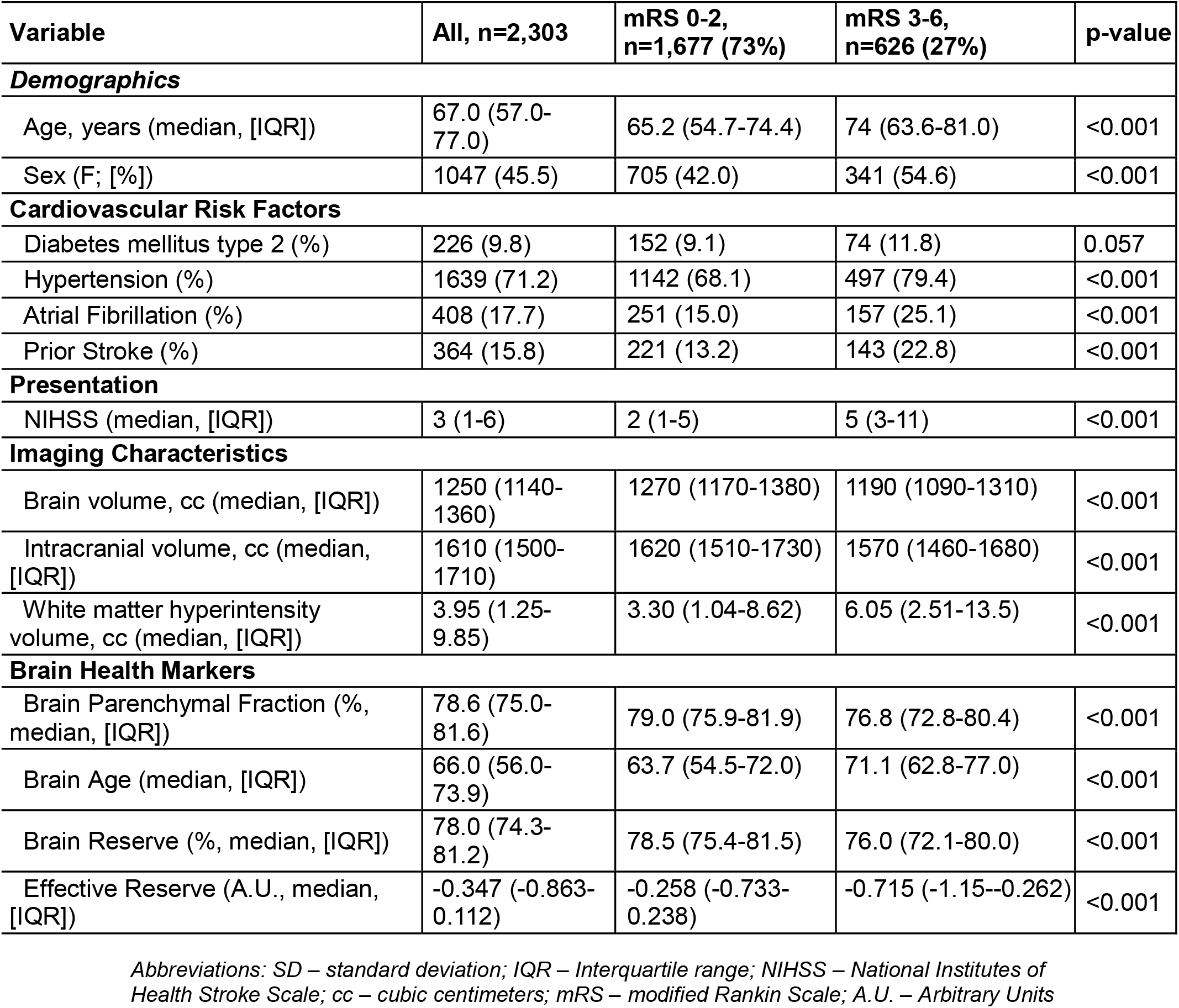
Characteristics of 2,303 Patients with Acute Ischemic Stroke.

All analyses were performed using R (version 4.4.1; R Foundation for Statistical Computing) with significance level set to p<0.05.

### Data Availability

Data, methods and materials will be made available for the purpose of reproducing the results upon reasonable request to the authors, subject to approval by the local Institutional Review Boards.

## Results

### Study Cohort and Baseline Characteristics

We analyzed 2,660 patients with complete baseline data (median age 67 years, 46% female), of which 357 (13.4%) were excluded due to missing follow-up data (see Figure 1). Baseline characteristics stratified by outcome categories appear in Table 1. Baseline characteristics included hypertension (71%), diabetes mellitus type 2 (10%), atrial fibrillation (18%) and history of prior stroke (16%). Median NIHSS score was 3 (IQR 1-6), median brain volume 1250 cc (IQR 1140-1360) and median WMH volume 3.95 cc (IQR 1.25-9.85). At 90 days, 626 patients (27%) had a mRS score of 3-6.

### Comparison of Model Performance with and without Brain Health Markers

Investigated models are presented with their corresponding BICs and PR-AUC values in Table 2 and graphically with adjusted odds ratios (aORs) and 95% CI in Figure 2.

**Table 2.** Comparison of Investigated Models Including a Quantitative MRI Marker of Brain Health to Predict Functional Outcome defined as mRS 3-6 at 90 Days After Acute Ischemic Stroke.

| Model | Included variables | aOR | 95% CI | P-value | BIC | PR-AUC (95% CI) |
| --- | --- | --- | --- | --- | --- | --- |
| Reference | Age | 1.04 | 1.03 - 1.05 | <0.001 | 2354.9 | 0.753 (0.668 - 0.827)<br>p-value: N/A |
|  | Female | 1.47 | 1.20 - 1.81 | <0.001 |  |  |
|  | Diabetes Type 2 | 1.32 | 0.96 - 1.82 | 0.087 |  |  |
|  | Hypertension | 1.30 | 1.02 - 1.70 | 0.037 |  |  |
|  | NIHSS | 1.15 | 1.13 - 1.17 | <0.001 |  |  |
|  | Atrial Fibrillation | 1.12 | 0.86 - 1.45 | 0.385 |  |  |
|  | Prior Stroke | 1.92 | 1.48 - 2.48 | <0.001 |  |  |
| Brain Parenchymal Fraction (BPF) | BPF, % | 0.96 | 0.94 - 0.98 | <0.001 | 2347.9 | 0.756 (0.669 - 0.833) p-value: 0.472 |
|  | Age | 1.03 | 1.02 - 1.04 | <0.001 |  |  |
|  | Female | 1.49 | 1.22 - 1.83 | <0.001 |  |  |
|  | Diabetes Type 2 | 1.38 | 0.99 - 1.90 | 0.053 |  |  |
|  | Hypertension | 1.26 | 0.99 - 1.62 | 0.066 |  |  |
|  | NIHSS | 1.15 | 1.13 - 1.18 | <0.001 |  |  |
|  | Atrial Fibrillation | 1.13 | 0.87 - 1.46 | 0.371 |  |  |
|  | Prior Stroke | 1.83 | 1.41 - 2.37 | <0.001 |  |  |
| Brain Age (BA) | BA | 1.05 | 1.04 - 1.06 | <0.001 | 2329.3 | 0.758 (0.688 - 0.833) p-value: 0.385 |
|  | Female | 1.59 | 1.29 - 1.95 | <0.001 |  |  |
|  | Diabetes Type 2 | 1.27 | 0.92 - 1.75 | 0.147 |  |  |
|  | Hypertension | 1.27 | 1.00 - 1.64 | 0.057 |  |  |
|  | NIHSS | 1.15 | 1.13 - 1.17 | <0.001 |  |  |
|  | Atrial Fibrillation | 1.17 | 0.90 - 1.51 | 0.23 |  |  |
|  | Prior Stroke | 1.81 | 1.39 - 2.34 | <0.001 |  |  |
| Brain Reserve (BR) | BR, % | 0.95 | 0.93 - 0.97 | <0.001 | 2343.7 | 0.757 (0.671- 0.833) p-value: 0.353 |
|  | Age | 1.03 | 1.02 - 1.04 | <0.001 |  |  |
|  | Female | 1.50 | 1.22 - 1.84 | <0.001 |  |  |
|  | Diabetes Type 2 | 1.36 | 0.98 - 1.87 | 0.064 |  |  |
|  | Hypertension | 1.25 | 0.98 - 1.61 | 0.077 |  |  |
|  | NIHSS | 1.15 | 1.13 - 1.17 | <0.001 |  |  |
|  | Atrial Fibrillation | 1.13 | 0.87 - 1.46 | 0.362 |  |  |
|  | Prior Stroke | 1.82 | 1.40 - 2.36 | <0.001 |  |  |
| effective Reserve (eR) | eR | 0.39 | 0.33 - 0.47 | <0.001 | 2318.4 | 0.763 (0.681 - 0.832)<br>p-value: 0.038 |
|  | Female | 1.06 | 0.85 - 1.31 | 0.626 |  |  |
|  | Diabetes Type 2 | 1.25 | 0.90 - 1.73 | 0.172 |  |  |
|  | Hypertension | 1.19 | 0.93 - 1.54 | 0.164 |  |  |
|  | NIHSS | 1.15 | 1.13 - 1.18 | <0.001 |  |  |
|  | Atrial Fibrillation | 1.19 | 0.92 - 1.53 | 0.193 |  |  |
|  | Prior Stroke | 1.74 | 1.34 - 2.25 | <0.001 |  |  |

*Abbreviations: aOR – adjusted Odds Ratio; BIC – Bayesian Information Criterion; CI – Confidence*
*Interval; PR-AUC – Area Under the Precision Recall Curve; NIHSS – National Institutes of Health*
*Stroke Scale; WMH – white matter hyperintensity; BPF – Brain Parenchymal Fraction; BA –*
*Radiomics derived Brain Age; BR – Brain Reserve; eR – effective Reserve.*

**Figure 2.**
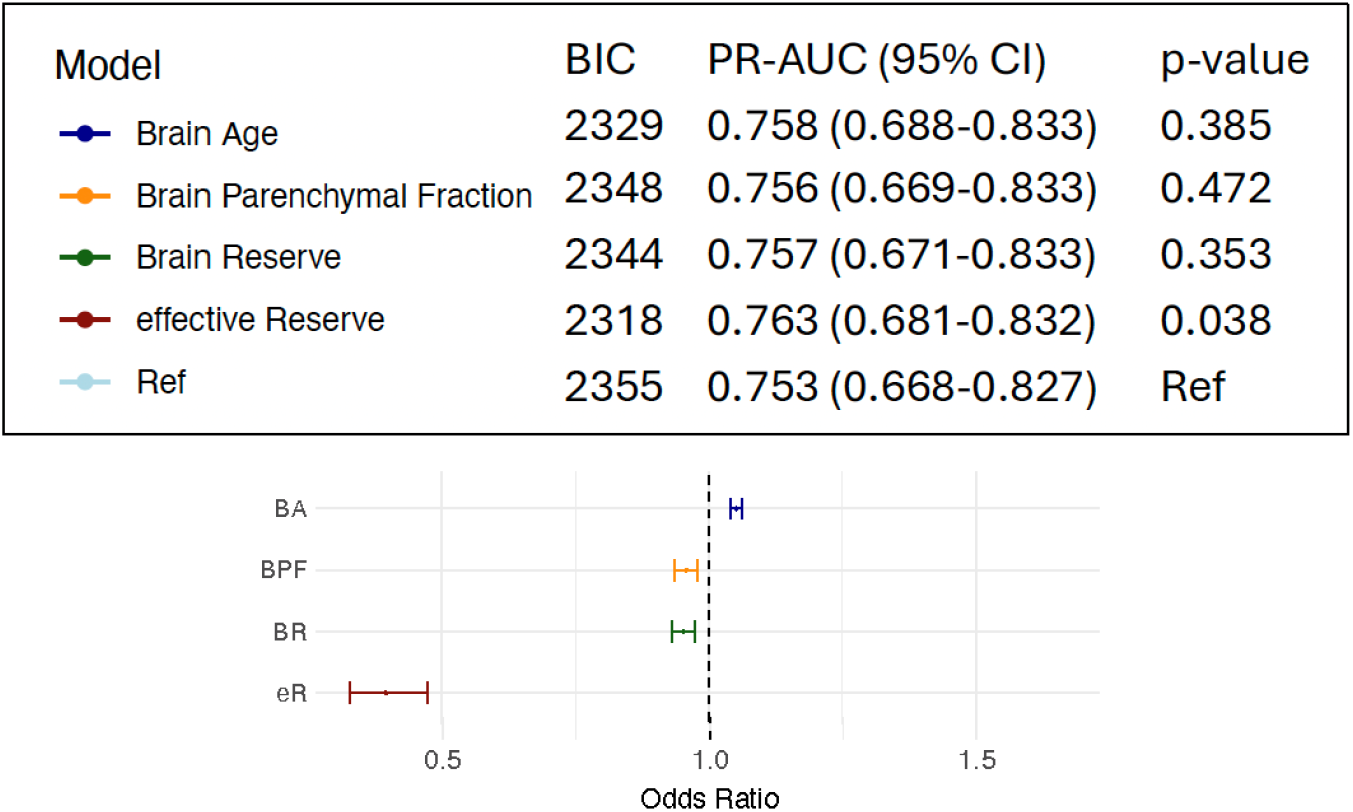
Comparison of Models to Predict Functional Outcome (mRS 3-6) at 90 days After Acute Ischemic Stroke. Abbreviations: BIC - Bayesian Information Criterion; PR-AUC – Precision Recall Area Under Curve; Ref – Reference; CI – Confidence Interval; BA – radiomics derived Brain Age; BPF – Brain Parenchymal Fraction; BR – Brain Reserve; eR – effective Reserve; NIHSS – National Institutes of Health Stroke Scale; mRS – modified Rankin Scale; DM – Diabetes Mellitus Type 2; HTN – Hypertension. Each brain health model and its adjusted odds ratio appear as a colored dot and corresponding 95% confidence interval line. Models were adjusted for age, sex, prior stroke, atrial fibrillation, NIHSS, hypertension, diabetes type 2 (see Table 2 for variables included in each model), Lower BIC indicate better model performance. In model selection analysis, evidence levels for statistical differences are considered Weak, Positive, Strong or Very strong for differences in BIC (ΔBIC) larger than 0, 2, 6 and 10 respectively. PR-AUC values for each model including a quantitative MRI marker of brain health are compared to a reference model with only clinical variables using Mann Whitney U test.

Comparison using BIC provided strong statistical evidence (ΔBIC > 6)^15^ for all four models including a brain health marker to outperform the reference model.

The model utilizing eR showed the lowest BIC value (BIC=2318.4), providing very strong (ΔBIC > 10) statistical evidence to outperform all other investigated models (see Table 2 and Figure 2).^15^ BA showed the second lowest BIC value (BIC=2329.3), providing very strong (ΔBIC > 10) statistical evidence to outperform all other models except the eR model. The PR-AUC value was significantly higher in the eR (0.763, 95% CI 0.681-0.832, p=0.038) compared to the reference model (0.753, 95% CI 0.668-0.827), but no significant differences were observed compared to the BA, BPF or BR models. VIF values indicated no significant multicollinearity for variables included in the investigated models respectively.

### Subanalysis with Comprehensive Model Adjustments

Results from the subanalysis are presented with the models’ corresponding BICs and PR-AUC values in eTable 1. Model comparison using BIC provided very strong statistical evidence for eR (BIC=2318.4, ΔBIC > 10) to outperform BPF (BIC=2334.9), BR (BIC=2343.7) and the reference models (BIC=2329.0) and strong statistical evidence to outperform the BA model (BIC=2324.9, ΔBIC > 2). The model utilizing BA outperformed the reference model (ΔBIC > 2) and the BPF and BR models (ΔBIC > 10). In the subanalysis, we observed no difference in PR-AUC values between any of the brain health models, as compared to the reference model.

## Discussion

In this international observational multicenter study, we demonstrate that the addition of neuroimaging brain health markers enhances modeling of functional outcome after AIS. The model utilizing effective Reserve (eR) outperformed the other investigated models, highlighting eR as a promising marker to quantify brain health in clinical settings.

Our findings are in line with previous studies showing the importance of incorporating brain health markers to predict outcomes after neurological diseases.^11-14,17^ Mostly smaller cohorts have explored the use of brain health markers specifically in stroke, often relying on high quality imaging data and manual imaging assessment.^11,17,18^ Here, we utilize automatized pipelines to quantify structural brain health, enabling translation into the routine acute stroke setting and bringing the holistic concept of *brain health in stroke* closer to clinical implementation. From a clinical perspective, such markers may complement established clinical and imaging predictors by providing an estimate of the brain’s capacity to tolerate ischemic injury or recovery potential, thereby refining individualized prognostic assessment in the acute and subacute stroke settings. While the methodology used to extract the neuroimaging markers are also applicable to modern ischemic stroke patient cohorts, prospective validation in contemporary cohorts is required before routine clinical implementation can be recommended.

Both eR and radiomics derived BA outperformed the other models, with substantially lower BICs and the eR model additionally showed higher PR-AUC, compared to the clinical reference model (Table 2). While statistically significant, the numerical differences in PR-AUC were relatively modest and implications for prognostication, risk stratification and trial designs needs to be investigated in future work. The results were consistent regarding BICs, but not PR-AUCs, when adjusting more comprehensively also for WMH load and brain volume (eTable 1). A possible explanation is that BPF and BR have not been specifically developed to quantify brain health in stroke populations and BR do not comprehend present pathology. The latent variable approach used to estimate eR and the radiomics approach to calculate BA may capture more nuanced signals of brain health, as compared to markers of brain volume, WMH volume and ICV, combined or alone.

While brain health is a multidimensional concept encompassing several domains, the current study focused on markers of structural brain health that are directly available at time of admission. This study provides evidence for associations between the investigated measures of brain health and functional outcome, however, causal relationships remain to be explored. Future studies are needed to determine whether brain health is an independent predictor of functional outcomes or, instead, a quantitative measure of resilience that reflects the capacity to mitigate the negative effects of brain injury.

Strengths of this study include the large international multicenter setting with standardized automated central adjudication of imaging to quantify structural brain health. Several limitations warrant comment. First, study participants were enrolled between 2003-2011, detailed treatment data were not available and included patients largely presented with mild to moderate stroke severity. The cohort may hence not be fully representative of severe stroke and current stroke care, involving more inclusive and advanced reperfusion therapies. The generalizability of our findings to contemporary stroke patient cohorts should be interpreted with caution. Second, investigated brain health markers are dependent on MRI which is not broadly available in acute stroke care worldwide. Third, only structural measures of brain health were considered, and we acknowledge that the limited available sequences in our data set did not allow for taking other imaging markers such as cerebral microbleeds, macrohemorrhages and measures of structural connectivity into account. Fourth, data on timing of stroke symptom onset, stroke phenotypes were limited, and brain health markers were assessed after stroke onset. Acute ischemic changes potentially influence the estimation of volumetric measures and WMH segmentation. However, the pipelines used have been developed in stroke patient cohorts and specifically designed not to misinterpret ischemic hyperintensities as WMH. Further, according to the study protocol, only patients with acute stroke were consecutively included at time of their acute hospital admission, supporting a restricted time between stroke onset and image assessment and our results to reflect mostly acute strokes.^6^ Fifth, data on premorbid mRS was not available. Instead, we included prior stroke as a proxy for premorbid status and adjusted for several comorbidities in the multivariate models. Cognitive aspects of brain health and a wider variety of outcomes, e.g. those obtained via patient reported outcome measures (PROMs), remain to be investigated. Next steps should further investigate how AIS or modifiable risk factors influence brain health and whether brain health modifies the outcome after stroke.

In conclusion, our results emphasize the importance of incorporating MRI measures of brain health for personalized outcome prognostication after AIS and suggest eR as a viable marker for future studies investigating brain health and stroke.

### Study Funding

Research reported in this publication was supported by the Swedish Research Council (2023-06531), the Swedish Heart Lung Foundation (no. 20230904, 2024135125), The Swedish Brain Foundation, The Swedish Government (under the “Avtal om Läkarutbildning och Medicinsk Forskning, ALF”), Lund University, Region Skåne, The FGS Fang Foundation, The Fremasons Lodge of Instruction Eos in Lund, The Swedish Stroke Association, AI Cures Grant Award, and the National Institute on Aging (NIA R21AG083559).

## Disclosure

NSR is supported by NINDS U19NS115388 and the MGB AI Cures Grant Award. AGL is national leader for Sweden for one ongoing stroke trial, local PI for the StrokeCLOSE study, Co-chairs the Global Alliance for International Stroke Genetics Consortium of Acute and Long-term Outcome studies, is Co-PI for The Genetics of Ischaemic Stroke Functional Outcome (GISCOME), and reports personal fees from Arega, Bayer, Astra Zeneca, BMS Pfizer, and Novo Nordisk. RWR serves on a DSMB for a trial sponsored by Rapid Medical, serves as site PI for studies sponsored by MicroVention and Penumbra, and receives research grant support from NIH (NINDS R25NS065743), Society of Vascular and Interventional Neurology, and Heitman Stroke Foundation. MDS is supported by AI Cures Grant Award, Heitman Stroke Foundation, and NIA R21AG083559.

## Acknowledgements

The authors thank participating patients and their families and the MRI-GENIE and GISCOME investigators.

Research reported in this publication was supported by the National Institute On Aging of the National Institutes of Health under Award Number R21AG083559. The content is solely the responsibility of the authors and does not necessarily represent the official views of the National Institutes of Health. This manuscript is the result of funding in whole or in part by the National Institutes of Health (NIH). It is subject to the NIH Public Access Policy. Through acceptance of this federal funding, NIH has been given a right to make this manuscript publicly available in PubMed Central upon the Official Date of Publication, as defined by NIH.

## Appendix

**MRI-GENIE Study Group Members**

| Name | Location | Role | Contribution |
| --- | --- | --- | --- |
| Daniel Woo, MD | Department of Neurology and Rehabilitation Medicine, University of Cincinnati College of Medicine, Cincinnati, OH, and Department of Neurology, Jacobs School of Medicine and Biomedical Sciences, University of Buffalo, NY, USA | Coinvestigator, Site PI | Data acquisition |
| Natalia S. Rost, MD, PhD | J. Philip Kistler Stroke Research Center, Massachusetts General Hospital, Harvard Medical School, Boston, MA, USA | Author, Site PI | Revision of the manuscript for content, including medical writing for content, Major role in the acquisition of data, analysis or interpretation of data |
| Jordi Jimenez-Conde, MD | Department of Neurology, Neurovascular Research Group (NEUVAS), IMIM Hospital del Mar (Institut Hospital del Mar d'Investigacions Mèdiques), Universitat Autònoma de Barcelona, Barcelona, Spain | Coinvestigator, Site PI | Data acquisition |
| Christina Jern, MD, PhD | Institute of Biomedicine, Department of Laboratory Medicine, the Sahlgrenska Academy, University of Gothenburg, Gothenburg, Sweden; Region Västra Götaland, Sahlgrenska University Hospital, Department of Clinical Genetics and Genomics, Gothenburg, Sweden | Author, Site PI | Revision of the manuscript for content, including medical writing for content, Major role in the acquisition of data, analysis or interpretation of data |
| Arne G. Lindgren, MD, PhD | Section of Neurology, Skåne University Hospital, Lund, Sweden; Department of Clinical Sciences Lund, Neurology, Lund University, Lund, Sweden | Author, Site PI | Revision of the manuscript for content, including medical writing for content, Major role in the acquisition of data, analysis or interpretation of data |
| Robin Lemmens, MD | KU Leuven - University of Leuven, Department of Neurosciences, Experimental Neurology and Leuven Research Institute for Neuroscience and Disease (LIND), Leuven, Belgium; VIB, Vesalius Research Center, Laboratory of Neurobiology, University Hospitals Leuven, Department of Neurology, Leuven, Belgium | Coinvestigator, Site PI | Data acquisition |
| Turgut Tatlisumak, MD, PhD | Department of Neurology, Helsinki University Hospital, Helsinki, Finland, Department of Clinical Neuroscience, Institute of Neuroscience and Physiology, Sahlgrenska Academy at University of Gothenburg, Gothenburg, Sweden; Department of Neurology, Sahlgrenska University Hospital, Gothenburg, Sweden | Coinvestigator, Site PI | Data acquisition |
| Ramin Zand, MD | Department of Neurology, Geisinger, Danville, PA | Coinvestigator, Site PI | Data acquisition |

GISCOME Study Group Members
| Name | Location | Role | Contribution |
| --- | --- | --- | --- |
| Christina Jern, MD, PhD | Institute of Biomedicine, Department of Laboratory Medicine, the Sahlgrenska Academy, University of Gothenburg, Gothenburg, Sweden; Region Västra Götaland, Sahlgrenska University Hospital, Department of Clinical Genetics and Genomics, Gothenburg, Sweden | Author, Study Group PI and representative | Revision of the manuscript for content, including medical writing for content, Major role in the acquisition of data, analysis or interpretation of data |
| Arne G. Lindgren, MD, PhD | Section of Neurology, Skåne University Hospital, Lund, Sweden; Department of Clinical Sciences Lund, Neurology, Lund University, Lund, Sweden | Author, Study Group PI and representative | Revision of the manuscript for content, including medical writing for content, Major role in the acquisition of data, analysis or interpretation of data |
| Jane Maguire, PhD, BA, BNurs (Hons) | University of Technology Sydney, Sydney, Australia | Author, Study Group PI and representative | Revision of the manuscript for content, including medical writing for content, Major role in the acquisition of data,<br>analysis or interpretation<br>of data |

## Notes

### Competing Interest Statement

The authors have declared no competing interest.

### Author Declarations

IRB of Massachusetts General Hospital gave ethical approval of this work, protocol numbers: GASROS #: 2001P001186; MRI-GENIE #: 2001P001186

### Summary of Updates

Updated Title, Abstract. Clarified and expanded Methods, Results and Discussion.

